# AENEAS Project: First real-time intraoperative application of machine vision-based anatomical guidance in neurosurgery

**DOI:** 10.64898/2026.04.09.26348607

**Authors:** Gary Sarwin, Vittorio Ricciuti, Victor E. Staartjes, Alessandro Carretta, Nadim Daher, Zhijin Li, Luca Regli, Diego Mazzatenta, Matteo Zoli, Ryu Seungjun, Ender Konukoglu, Carlo Serra

## Abstract

**Background and Objectives:** We report the first intraoperative deployment of a real-time machine vision system in neurosurgery, derived from our previous anatomical detection work, automatically identifying structures during endoscopic endonasal surgery. Existing systems demonstrate promising performance in offline anatomical recognition, yet so far none have been implemented during live operations.

**Methods:** A real-time anatomy detection model was trained using the YOLOv8 architecture (Ultralytics). Following training completion in the PyTorch environment, the model was exported to ONNX format and further optimized using the NVIDIA TensorRT engine. Deployment was carried out using the NVIDIA Holoscan SDK, the system ran on an NVIDIA Clara AGX developer kit. We used the model for real-time recognition of intraoperative anatomical structures and compared it with the same video labelled manually as reference. Model performance was reported using the average precision at an intersection-over-union threshold of 0.5 (AP50). Furthermore, end-to-end delay from frame acquisition to the display of the annotated output was measured.

**Results:** A mean AP50 of 0.56 was achieved. The model demonstrated reliable detection of the most relevant landmarks in the transsphenoidal corridor. The mean end-to-end latency of the model was 47.81 ms (median 46.57 ms).

**Conclusion:** For the first time, we demonstrate that clinical-grade, real-time machine-vision assistance during neurosurgery is feasible and can provide continuous, automated anatomical guidance from the surgical field. This approach may enhance intraoperative orientation, reduce cognitive load, and offer a powerful tool for surgical training. These findings represent an initial step toward integrating real-time AI support into routine neurosurgical workflows.

## INTRODUCTION

Successful outcomes in neurosurgery do not rely only on technical skills, but also on cognitive mastery of the procedure. While surgical dexterity can be improved through training, or ideally even robotic assistance, the real challenge often lies in the surgeon’s mental roadmap, which can be defined as the internalized plan that guides each step of the procedure. Unlike technical skills, the mental roadmap is heavily dependent on spatial reasoning, expertise, and on exposure to valid mentors and peers during the years. In complex neurosurgical procedures, the variability in cognitive navigation is a source of inter-surgeon difference and operative risk that remains challenging to reduce.

Over the past decades, several technologies have been developed to support intraoperative guidance and orientation, such as neuronavigation, intraoperative ultrasound, intraoperative MRI, 5-ALA, intraoperative neuromonitoring^1–5^. Despite their undoubted usefulness, none of these provide real-time, same-modality picture-in-picture information, and carry well-known modality-specific disadvantages (operator-dependent interpretation, additional infrastructure costs, aspecificity, etc.)^6,7^. Consequently, the burden of constructing and maintaining the mental roadmap remains largely with the surgeon.

Artificial intelligence and machine/computer vision offer a promising strategy to augment this cognitive process. In our initial proof-of-concept studies, we demonstrated that deep learning algorithms could detect key anatomical structures from pre-recorded endoscopic or microscopic videos^8,9^, establishing the feasibility of automated landmark recognition. Furthermore, we were able to expand this capability by integrating detection into a roadmap-based framework, enabling the encoding of spatial relationships among anatomical structures^10,11^. This approach was applied not only to endoscopic transsphenoidal procedures but also to open neurosurgical approaches, including the pterional transsylvian approach, where detection of deep structures such as the optic nerve and internal carotid artery proved feasible^12,13^. Collectively, these studies show that machine vision can replicate components of the surgeon’s mental roadmap across different neurosurgical contexts.

Here, we report the first application of the AENEAS Project framework for real-time anatomical recognition during live endoscopic procedures, demonstrating that machine vision could support intraoperative decision-making and enhance the surgeon’s mental roadmap. By translating previous advances from retrospective video analysis into the live surgical setting, this work represents a critical step toward the clinical integration of AI-driven cognitive guidance in neurosurgery.

## METHODS

### AI Model Development and Training

A real-time object-detection model was developed using the YOLOv8 architecture (Ultralytics)^14^, selected for its established performance in high-throughput computer-vision applications. The model was trained for 147 epochs. To ensure that model performance reflected true generalizability rather than memorization of patient-specific features, we enforced strict patient-level separation. All frames originating from a single patient were allocated exclusively to one of the dataset splits (training, validation, or test). The final model was selected on the basis of validation-set performance and was subsequently evaluated on the completely independent test set.

Model performance was reported using the average precision at an intersection-over-union threshold of 0.5 (AP50), in accordance with accepted standards in real-time object detection. Class-specific AP50 values are summarized in Table 1. Performance during the live neurosurgical case was assessed retrospectively by frame-by-frame analysis, as described below.

**Table 1.** Test set showing class performances of the object detection model.

| Class | Precision | Recall | AP50 |
| --- | --- | --- | --- |
| Nasal Septum | 0.596 | 0.835 | 0.801 |
| SupM | 0.277 | 0.508 | 0.305 |
| MidM | 0.468 | 0.737 | 0.659 |
| InfM | 0.459 | 0.677 | 0.548 |
| Coana | 0.489 | 0.561 | 0.516 |
| Floor | 0.556 | 0.643 | 0.685 |
| RecSphEthm | 0.399 | 0.386 | 0.318 |
| Sphenoid Ostium | 0.536 | 0.495 | 0.486 |
| Instrument | 0.892 | 0.963 | 0.971 |
| Rostrum | 0.070 | 0.056 | 0.045 |
| Sphenoidal Sinus | 0.56 | 0.860 | 0.753 |
| Sella Floor | 0.62 | 0.646 | 0.640 |
| Clival Recess | 0.405 | 0.549 | 0.438 |
| Planum | 0.319 | 0.525 | 0.315 |
| Left Osseous Carotis | 0.395 | 0.228 | 0.231 |
| Right Osseous Carotis | 0.266 | 0.235 | 0.152 |

*Abbreviations: SupM, Superior Nasal Meatus; MidM, Middle Nasal Meatus; InfM, Inferior Nasal Meatus; RecSphEthm, Sphenoethmoidal Recess; AP50, average precision at an intersection-over-union threshold of 0.5.*

### Dataset

The study protocol was reviewed and approved by the local Ethics Committee (KEK 2023-02265), and all procedures were performed in accordance with the ethical standards laid down in the Declaration of Helsinki.

### Source Data and Consent

The training data consisted of anonymised surgical videos obtained from patients undergoing endoscopic transsphenoidal surgery for pituitary adenoma resection. The dataset originates from 169 individual operations performed using a variety of rigid endoscopes over a ten-year period at several neurosurgical centers within the same geographic region. The participants and any identifiable individuals consented to publication of his/her image. Prior to inclusion, each video was anonymised by the removal of metadata and any potential indirect identifiers.

### Video Acquisition and Annotation

For the present study, we used an annotated dataset with a total of 22 584 labelled frames from 169 patients. Each frame was labelled manually by neurosurgeons, who identified all visible instances of sixteen predefined classes: fifteen anatomical structures of relevance to the transsphenoidal route and one dynamic surgical-instrument class. Owing to the fixed nature of skull-base anatomy, each image contained at most one instance of any anatomical class, whereas the instrument class could appear multiple times across frames.

### Dataset Composition

Of the 169 patients represented in the expanded dataset, 153 were allocated to the training set, eight to the validation set, and eight to the independent test set. This resulted in 22 584 annotated frames and a total of 62819 bounding-box annotations. The distribution of annotations was strongly class-imbalanced, with the most common class (“Instrument”) accounting for 14 682 annotations and the least frequent (“Floor”) accounting for 684 annotations, corresponding to a 21.5:1 imbalance ratio. The class distribution is visualised in Figure 1.

**Figure 1.**
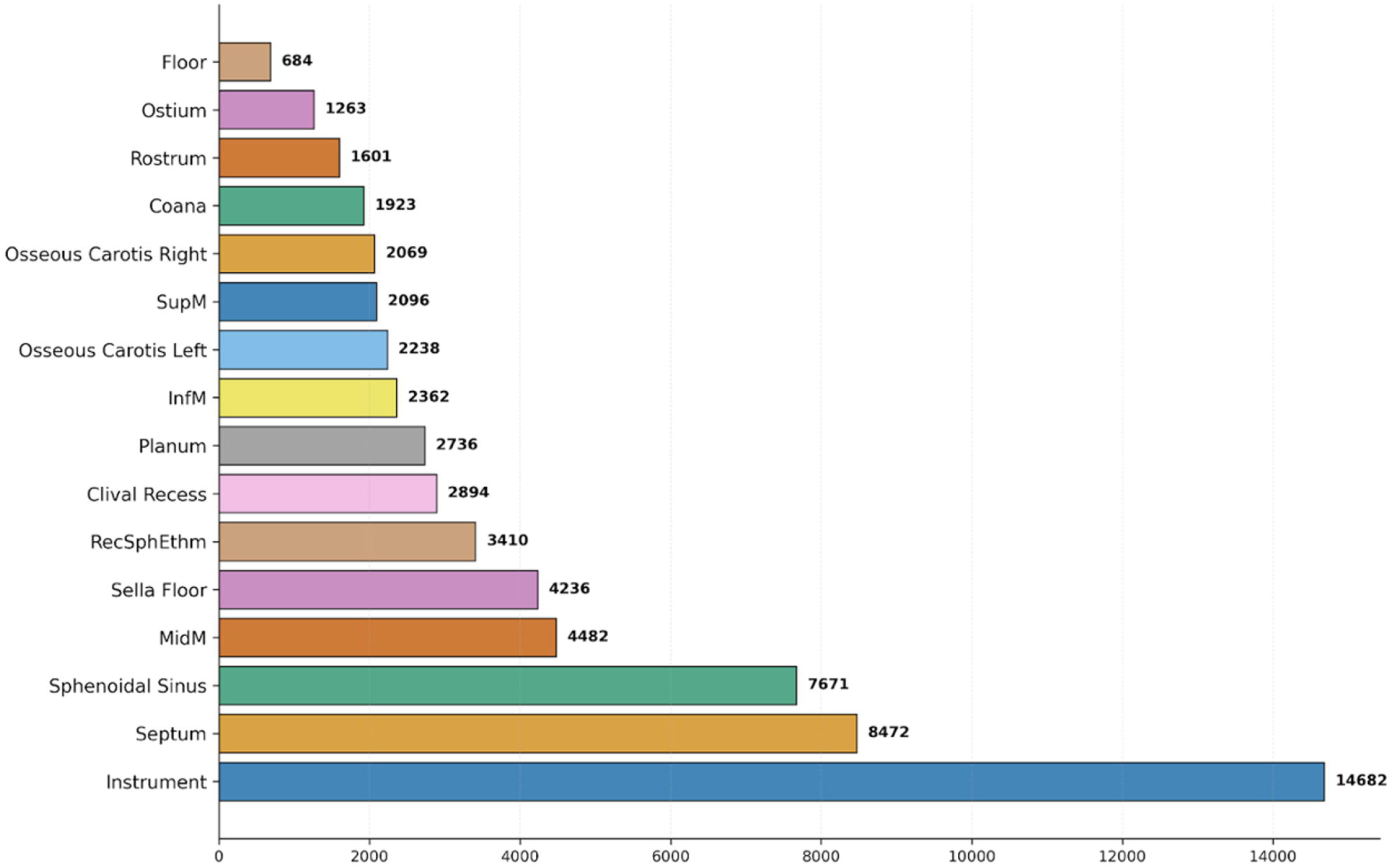
Overall Class Distribution: in the histogram are reported the number of the annotations for each class (i.e. anatomical structure) manually labelled in the video database used for the training set.

### Model Optimization and Deployment

Following completion of training in the PyTorch environment^15^, the model was exported to ONNX format^16^ and further optimized using the NVIDIA TensorRT engine^17^.

### Software and Hardware Integration

Deployment was carried out using the NVIDIA Holoscan SDK^18^, which managed endoscopic video acquisition, preprocessing, model inference, and post-processing within a unified, low-latency computational graph. The system ran on an NVIDIA Clara AGX developer kit^19^ configured for real-time surgical imaging. The intraoperative endoscopic stream was delivered via SDI output from the endoscope, routed through a capture interface, and ingested into the Clara AGX. Each incoming frame underwent GPU-based preprocessing, followed by inference with the TensorRT-optimized YOLOv8 engine and post-processing steps including non-maximum suppression, and real-time overlay of anatomical labels. The annotated output was transmitted from the Clara AGX via DisplayPort to a separate dedicated monitor positioned within the operating room. For recording and subsequent analysis, the same DisplayPort signal was duplicated and sent to an additional capture card, enabling simultaneous capture of the processed video stream. Intraoperative set-up is shown in Figure 2.

**Figure 2.**
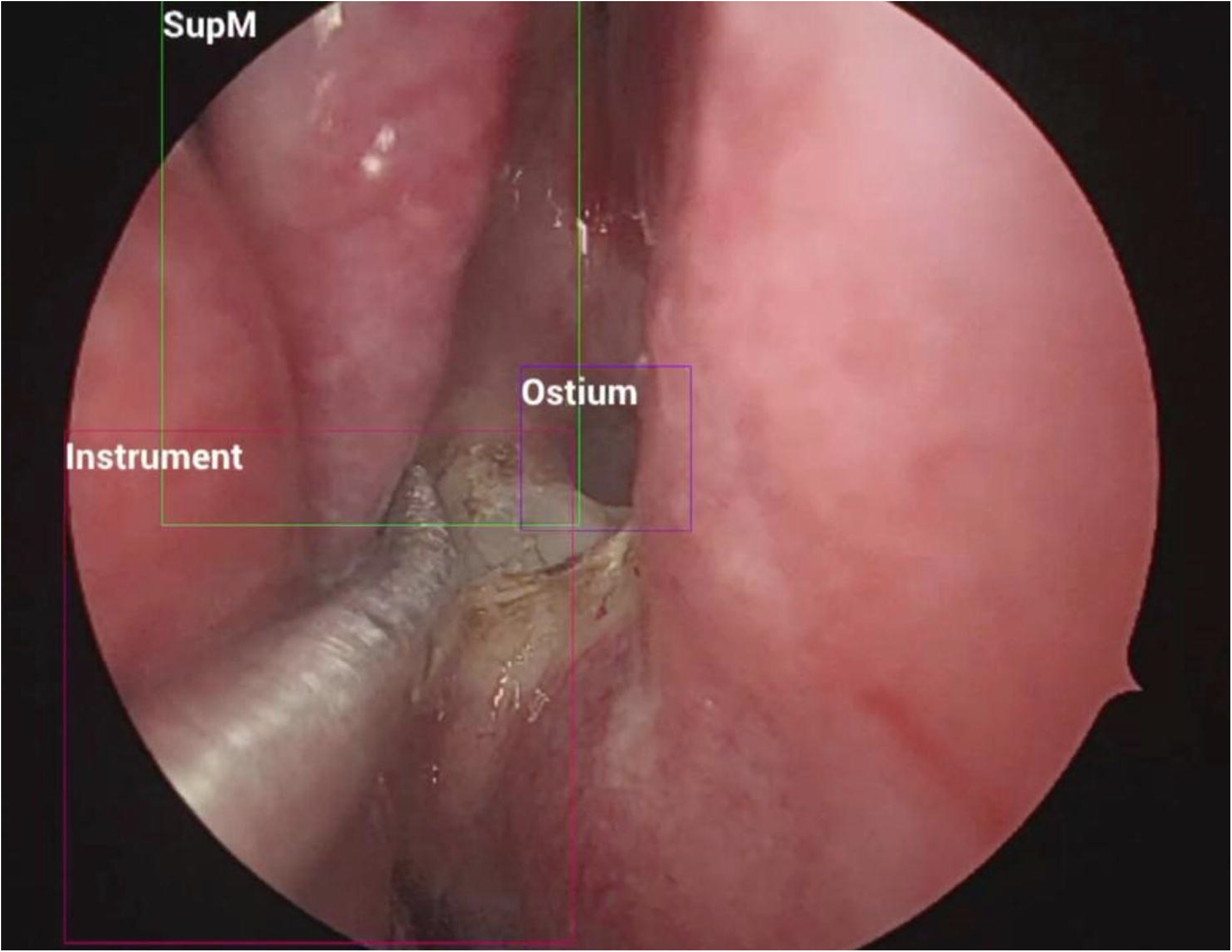
Operating room set-up: A) shows the additional monitor view, with the same video seen by the surgeon during surgery, but also with intraoperative automatic recognition of anatomical structures; B) shows the operating room set up, using the NVIDIA Holoscan SDK, equipped with the monitor used for displaying the annotated output. (B will be included in the final manuscript)

## RESULTS

### Model Performance on the Independent Test Set

The model demonstrated robust performance for the majority of the 16 classes in the test set. The mean AP50 was 0.491. Class-specific AP50 values are shown in Table 1 and range from 0.801 to 0.045 for the anatomical classes. Despite the class imbalance in the dataset, some structures such as “Floor” and “Ostium” and “Coana” still achieve promising results. Overall, the model demonstrated reliable detection of the most clinically relevant landmarks for navigation in the transsphenoidal corridor.

### Real-Time System Latency

A dedicated latency analysis was performed to characterize the performance of the real-time inference pipeline under operative-equivalent conditions. This assessment was conducted independently of the live surgical case using the built-in Holoscan latency profiler, which measures the full end-to-end delay from frame acquisition to the display of the annotated output. A sequence of 1000 consecutive frames was processed using the same pipeline configuration, TensorRT engine, and NVIDIA AGX platform used intraoperatively. The system exhibited stable and reproducible behavior. The mean end-to-end latency was 47.81 ms, with a median latency of 46.57 ms. The fastest frame was processed in 40.49 ms and the slowest in 72.06 ms, resulting in an effective throughput of approximately 21 frames per second. Ninety-nine percent of frames were processed within 66.20 ms, indicating minimal jitter and a consistent processing profile across the evaluation. This level of performance is compatible with intraoperative requirements, allowing smooth visualization and synchronized overlay of anatomical labels on the live endoscopic feed. As observed in similar real-time computer-vision systems, model inference time accounted for the majority of the total delay. Consequently, additional optimization strategies may further reduce latency. Moreover, next-generation embedded platforms such as the new NVIDIA IGX, which offers substantially higher real-time compute throughput and medical-grade reliability, are expected to reduce latency beyond what is achievable on the current AGX hardware, potentially enabling frame rates approaching native endoscopic video.

### Real-Time Detection Performance

Real-time detection performance during the live surgical case was assessed by retrospectively labelling 128 frames of the recorded surgical video and re-evaluating them using the PyTorch model. This yielded a mean AP50 of 0.56, consistent with the results observed in the independent test set. The “Floor” class is absent from this analysis, as it was not visible and therefore had no ground-truth annotations. Despite the limited number of frames, these findings confirm that real-time deployment did not materially degrade model performance. The full results are visualised alongside the Precision-Recall Curve in Figure 3.

**Figure 3.**
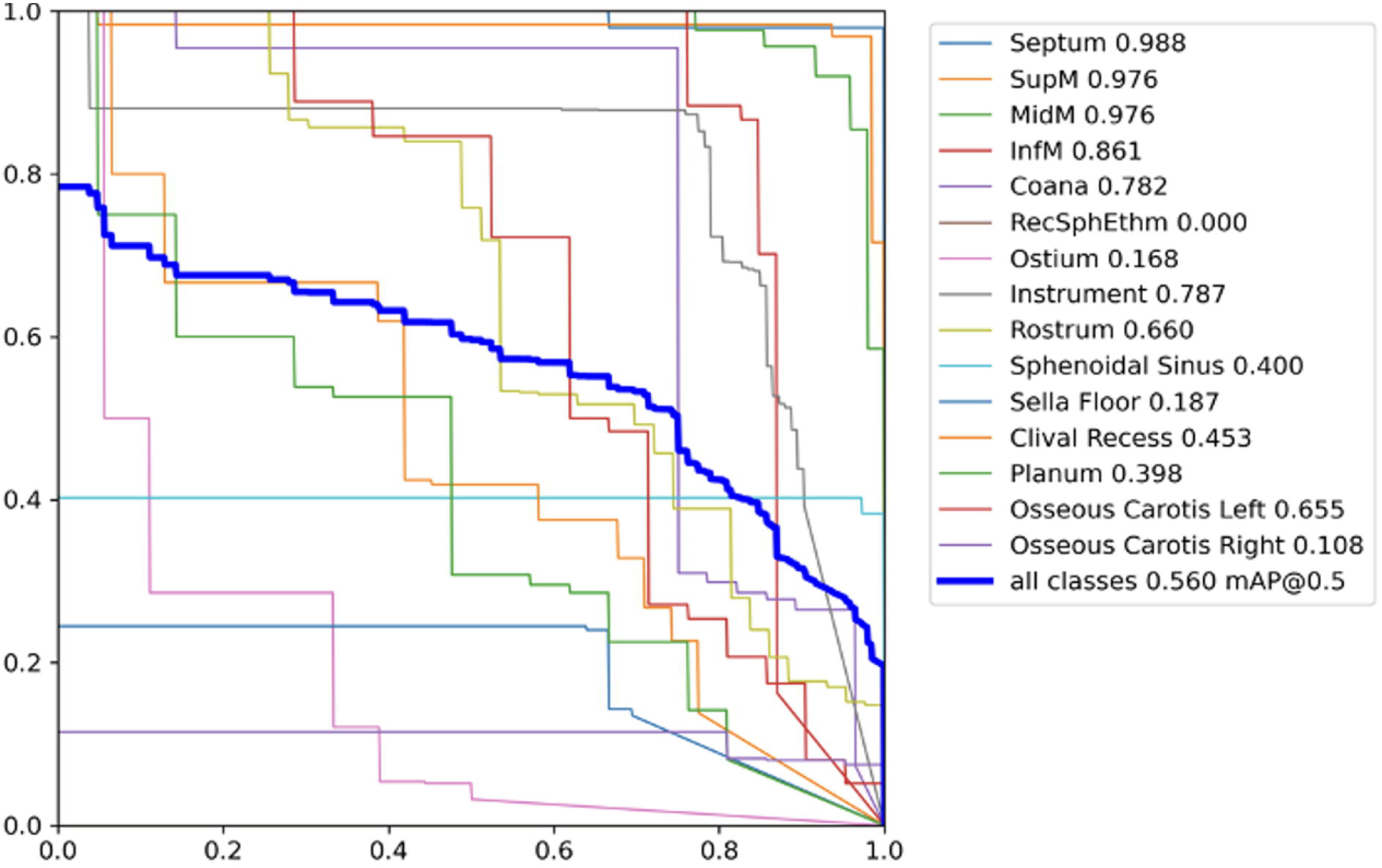
Real-time detection performance during live surgical case. Each point on the precision–recall curve represents a different detection confidence threshold (precision values on the y axis and recall values on the x axis). This figure shows how sensitivity to true findings (recall) increases as the rate of false positives rises (lower precision).

### Deployment

We demonstrate the deployment of AENEAS as a real-time tool for live endoscopic pituitary surgery in Video 1 (Set-Up) and Video 2 (Intraoperative Performance)

## DISCUSSION

The present study demonstrates, for the first time, the feasibility of real-time intraoperative anatomical recognition during endoscopic pituitary surgery. By deploying a fully optimized, low-latency machine vision pipeline directly utilizing the operative video stream, we show that automated detection of key anatomical structures is technically achievable and clinically compatible with the workflow of transsphenoidal surgery. The system consistently processed video frames within an average of 47.8 ms, a performance profile that ensured smooth visualization without perceptible delay. Importantly, real-time recognition accuracy remained aligned with the results obtained on the independent test set (mean live AP50 = 0.56), confirming that deployment did not degrade model performance.

### Clinical and Educational Implications of Real-Time Anatomical AI Guidance

This work represents a substantial advance over all prior machine vision studies in neurosurgery, which were limited to retrospective analyses or simulated intraoperative environments^9,12,13,20^. In contrast, our system operated live, identifying anatomical structures as they appeared during the surgical progression and effectively externalizing part of the surgeon’s cognitive roadmap^8,21^. By translating complex spatial reasoning into an automated, dynamic visual guide, this workflow can enhance intraoperative orientation within the narrow and shifting corridors of endoscopic skull-base surgery, where transient misrecognition of critical landmarks may increase operative risk. The model reliably recognized the most clinically relevant structures, including the septum, sphenoid sinus, sella floor, and sphenoid rostrum, despite class imbalance and variability in visualization; even for more challenging structures such as carotid protuberances, predictions were stable under optimal exposure. These results underscore the potential for performance gains through larger, more balanced datasets and expanded multimodal annotation strategies.

A variety of intraoperative imaging technologies have been developed to enhance surgical precision and situational awareness, each offering specific advantages but also important constraints. Preoperative navigation provides an initial anatomical roadmap, yet its reliability progressively declines as brain shift occurs during the procedure^22^. Ultrasound can offer real-time updates, although its effectiveness remains highly operator-dependent and its spatial resolution and interpretability can be inconsistent^23^. Intraoperative MRI ensures high-quality imaging but requires substantial resources, and often depends on dedicated infrastructures that limit its routine adoption^24^. Fluorescence-based techniques, while valuable for highlighting pathological tissue, are restricted to structures exhibiting specific uptake and therefore cannot deliver a comprehensive anatomical overview^25^. Finally, intraoperative neurophysiological monitoring and awake surgery paradigms further contribute by enabling real-time functional assessment during critical surgical steps; however, they rely on specialized technologies and highly trained personnel^26^.

Against this backdrop of useful yet fragmented or modality-specific technologies, our system introduces continuous, real-time anatomical information extracted directly from the operative video field. By integrating guidance seamlessly within the surgeon’s natural visual stream, it reduces the cognitive burden associated with reconciling disparate sources of information and avoids the need to shift attention away from the surgical scene. This enhanced immediacy not only supports more fluid and intuitive intraoperative decision-making but also has the potential to improve situational awareness and foster a more streamlined and efficient operative workflow.

Beyond its immediate clinical implications, this technology offers significant educational value: real-time anatomical recognition provides residents and fellows with continuous, objective cues during surgery, potentially accelerating acquisition of spatial understanding and standardizing intraoperative orientation across levels of experience^8,21,27^. In this sense, AI-driven anatomical detection functions as a form of virtual mentorship and may contribute not only to safer procedures but also to democratizing access to complex neurosurgical expertise^28,29^.

### Technical limitations and future perspectives

While the results are encouraging, some limitations remain. The current model is at this very moment optimized for transsphenoidal endoscopic surgery and has yet to be validated intraoperatively for other approaches or in the presence of significant pathological alterations. Nevertheless, the success of live implementation suggests that with larger, more diverse datasets, integration of additional anatomical structures, and continued refinement of the algorithm, broader application is achievable. Future developments may include mixed-reality integration, where anatomical recognition and roadmap projections are seamlessly overlaid on the surgeon’s visual field, with the option for on-demand activation. This could streamline navigation, enhance situational awareness, and improve workflow efficiency, without distracting from the operative scene. Furthermore, as the system matures, it could extend beyond guidance to the automation of complex procedural steps, modelling surgical expertise and making advanced techniques accessible to a wider range of practitioners.

## CONCLUSION

This study presents the first real-time, intraoperative use of machine vision for anatomical recognition in neurosurgery, showing that key structures can be automatically identified directly from operative video without relying on preoperative imaging or external tracking. This shift toward dynamic, real-time visual guidance may enhance surgical safety and precision, while potentially improving workflow efficiency and reducing cognitive load.

At the same time, real-time anatomical detection can offer valuable educational opportunities by supporting trainees and helping to standardize intraoperative orientation across operators. Future developments may broaden its anatomical and clinical applicability, validate its robustness in diverse scenarios, and enable integration with mixed-reality platforms, positioning machine vision navigation as a potentially transformative adjunct in neurosurgical practice.

## Data Availability

The data used is not publicly available.

## Acknowledgments

The authors wish to thank NVIDIA for their collaboration.

## Abbreviations

SupM: Superior Nasal Meatus
MidM: Middle Nasal Meatus
InfM: Inferior Nasal Meatus
RecSphEthm: Sphenoethmoidal Recess
AP50: average precision at an intersection-over-union threshold of 0.5.

## Notes

**Disclosures:** Some authors are employees of NVIDIA. The company provided hardware support used in this study. No other personal or financial conflicts of interest are declared. NVIDIA had no role in the design, execution, analysis, or reporting of the study.

### Competing Interest Statement

Some authors are employees of NVIDIA. The company provided hardware support used in this study. No other personal or financial conflicts of interest are declared. NVIDIA had no role in the design, execution, analysis, or reporting of the study.

### Funding Statement

This research is supported by SNF Project IZKSZ3_218786 research grant (Swiss National Funds)

### Author Declarations

Ethikkommission of Zurich, Switzerland gave ethical approval for this work (KEK 2023-02265)

